# Longitudinal associations among post-displacement stressors, physical activity, and mental health in Farsi- and Dari-speaking refugees and asylum-seekers in Australia

**DOI:** 10.64898/2026.08.11.26360157

**Authors:** G. Kurt, R. Rostami, G. McKeon, S. Rosenbaum, J. Solaimani, D. Berle, D. Silove, D. Hadzi-Pavlovic, Z. Steel, R. Wells

**Affiliations:** Discipline of Psychiatry and Mental Health, School of Clinical Medicine, Faculty of Medicine and Health, UNSW, Sydney; School of Population Health, Faculty of Medicine and Health, UNSW, Sydney; Institute of Physical Activity and Nutrition, School of Exercise and Nutrition Sciences, Deakin University, Melbourne, Australia; School of Medicine and Psychology, The Australian National University, Canberra, Australia; Centre for Mental Health and Brain Sciences, Swinburne University of Technology, Melbourne, Australia

**Keywords:** refugees, physical activity, post-displacement stressors, mental health

## Abstract

**Background:** Post-displacement stressors affect mental health among refugees and asylum-seekers, yet the behavioral mechanisms underpinning this relationship remain understudied.

**Objective:** To examine the role of physical activity in the relationship between post-displacement stressors and mental health outcomes among Farsi/Dari speaking refugees and asylum-seekers in Australia.

**Methods:** Data were drawn from a longitudinal community-based cohort study of 343 Farsi and Dari speaking refugees and asylum-seekers (80 female, 23.3%) in Australia conducted between 2017 and 2019. Data from post-displacement stressors measured at the baseline, physical activity at one-year follow-up, and mental health outcomes (symptoms of posttraumatic stress disorder (PTSD) and depression and personal mastery) at two-year follow-up were included in the study. Longitudinal path analyses were conducted to test the mediating role of moderate-to-vigorous physical activity and sedentary behaviour in the associations between post-displacement stressors and mental health outcomes.

**Results:** After controlling for baseline levels of depression and PTSD, traumatic events, and key demographic characteristics, post-displacement stressors significantly predicted less moderate-to-vigorous physical activity (MVPA) (β = −0.18, 95% CI [−0.292, −0.056]) and more sedentary behavior (β = 0.13, 95% CI [0.011, 0.243]) at one-year follow-up. Less MVPA, in turn, significantly predicted greater symptoms of depression (β = −0.21, 95% CI [−0.353, −0.060]) and lower level of personal mastery (β = 0.19, 95% CI [0.033, 0.327]) while greater time spent in sedentary behavior predicted greater symptoms of depression (β = 0.21, 95% CI [0.060, 0.359]), PTSD (β = 0.24, 95% CI [0.096, 0.386], and lower personal mastery ((β = −0.18, 95% CI [−0.336, −0.011]). Significant indirect associations were observed between post-displacement stressors and depressive symptoms and personal mastery through MVPA, and between post-displacement stressors and depressive and PTSD symptoms through sedentary behaviour.

**Conclusion:** These findings provide the first longitudinal evidence that moderate-to-vigorous physical activity and sedentary behavior partially explain the relationship between post-displacement stressors and subsequent mental health outcomes among refugees and asylum-seekers. Addressing these modifiable behaviors may represent targets for future intervention research to promote mental health during resettlement.

## Introduction

Humanitarian crises worldwide have led to the displacement of over 110 million individuals. There are currently over 40 million refugees and asylum-seekers who mostly live in low- and middle-income countries (United Nations High Commissioner for Refugees, 2025). Exposure to traumatic events and ongoing post-displacement stressors places forcibly displaced people at elevated risk of mental health problems and impaired functioning (Hou et al., 2020; Steel et al., 2009). Recent meta-analytic evidence demonstrates that one in three forcibly displaced people report posttraumatic stress disorder (PTSD) and depression (Blackmore et al., 2020). These estimates are almost five times higher than those observed in general population (GBD 2019 Mental Disorders Collaborators, 2022; Koenen et al., 2017), highlighting mental health problems associated with forced displacement as a growing public health concern. It is thus important to identify modifiable factors contributing to mental health burden in this population to guide development of targeted interventions and treatments.

Post-displacement conditions constitute key determinants of mental health following forced displacement. Numerous studies conducted across diverse resettlement and transit settings demonstrated that experiencing stressors such as legal insecurity, financial difficulties, housing problems, language barriers, and social isolation are associated with elevated levels of psychological distress, poor social functioning and diminished wellbeing (Hou et al., 2020). Recent research has increasingly examined the mechanisms through which post-displacement stressors contribute to poor mental health. To date, this work has largely focused on psychological mechanisms such as cognitive appraisals, attachment styles, and emotion-regulation. For instance, in a cross-sectional study among treatment-seeking people from refugee backgrounds in Switzerland, Nickerson et al. (2015) found that post-displacement stressors were associated with greater difficulties in emotion regulation, which in turn, predicted higher levels of depression and PTSD symptoms. Using the same dataset, Kurath et al. (2024) further showed that attachment styles, especially attachment anxiety, play a significant role in the relationship between post-displacement stressors and PTSD symptoms. In a longitudinal study of culturally and linguistically diverse community sample of refugees in Australia, post-displacement stressors predicted increases in emotion-regulation difficulties over time while emotion-regulation difficulties and post-displacement stressors demonstrated a bidirectional relationship and both predicted subsequent PTSD symptoms (Specker et al., 2024). Further, studies have shown that experiencing post-displacement stressors, particularly, lengthy asylum-seeking process, detention, fear of deportation might give rise to certain cognitive appraisals such as moral injury, defined as appraisals of events and/or actions as transgressing one’s moral beliefs, which are associated with poorer mental health (Donovan et al., 2025). While these findings provide important insights into potential intervention targets for attenuating the adverse effects of post-displacement stressors on mental health among refugees, this literature remains in its infancy. In particular, behavioral mechanisms have received little attention despite representing potentially modifiable targets that may complement existing psychological approaches and be readily translated into scalable interventions.

Physical activity is one potential behavioral pathway linking post-displacement stressors with mental health among refugees and asylum seekers. Broadly defined as “*moving, acting, and performing within culturally specific spaces and contexts*” (Piggin, 2020), physical activity encompasses movement undertaken across everyday life, including for transport, work, recreation and social participation. Meta-analytical evidence indicates that physical activity interventions can reduce symptoms of depression, anxiety, and PTSD across diverse populations (Noetel et al., 2024; Ramos-Sanchez et al., 2021; Rosenbaum et al., 2015), while prospective meta-analyses further suggest that regular physical activity is associated with a lower risk of developing depression and anxiety (Schuch et al., 2018, 2019). Greater sedentary behavior is also associated with poorer mental health, independent of physical activity (Allen et al., 2019; Schuch et al., 2017). Similar relationships between physical activity and mental health have been observed among trauma-exposed populations including survivors of natural disasters and veterans (Wang et al., 2023).

These behavioural pathways may be particularly relevant following displacement. Post-displacement conditions may restrict opportunities for physical activity and increase sedentary time through financial hardship, unsafe or unfamiliar environments, competing demands, social isolation and limited access to culturally appropriate activities (Hartley et al., 2017; McKeon et al., 2025). Lower physical activity and greater sedentary time may, in turn, contribute to poorer mental health through overlapping psychological, social and physiological pathways. Establishing whether these behaviours form part of the pathway between post-displacement stressors and mental health may therefore have important implications for prevention and care. Prior research conducted with refugee and asylum-seeker populations accord with evidence from general and other trauma-exposed populations. For instance, cross-sectional studies conducted in Sweden and Greece demonstrated that greater physical activity is associated with fewer PTSD symptoms, better wellbeing, lower stress, and improved sleep, whereas greater sedentary behavior is associated with poorer mental health among refugees (Filippou et al., 2024, 2025; Nilsson et al., 2019; Sjögren Forss et al., 2021). Although one study found that perceived physical fitness, rather than self-reported physical activity, was more consistently associated with mental health outcomes among refugees in a camp in Greece (Filippou et al., 2024), another study using objectively measured physical activity demonstrated that greater moderate-to-vigorous physical activity and lower sedentary behavior were associated with better mental health and wellbeing in the same setting (Filippou et al., 2025). In a community sample of refugees in Australia, Rostami et al. (2025) found that refugees and asylum seekers with insecure visa status were less likely to meet the World Health Organization’s physical activity guideline of 150 minutes per week and reported greater sedentary behavior than those with secure visas. Depression and PTSD symptoms mediated these associations, suggesting that mental health may also contribute to reduced physical activity and increased sedentary behavior. Together, these findings suggest that physical activity and mental health are closely linked among people from refugee backgrounds too. However, existing evidence is predominantly cross-sectional and no study to date investigated the mechanistic role of physical activity through which post-displacement stressors predict mental health among refugees.

The present study aimed to longitudinally examine the role of physical activity in the relationship between post-displacement stressors and mental health outcomes, namely, symptoms of depression and PTSD and personal mastery, in a community sample of Farsi speaking refugees and asylum-seekers in Australia. Australia is a major resettlement country in the Asia-Pacific region with one million refugees resettled since World War II. The majority of refugees in Australia come from Arabic and Farsi speaking backgrounds, the latter representing the largest group resettled through Australia’s offshore humanitarian program in the past five years (Department of Home Affairs, 2024, 2026; Minister for Foreign Affairs, 2026). Escalating humanitarian crisis in Afghanistan and ongoing civil unrest in Iran contributed to increasing numbers of forcibly displaced people from these countries seeking protection in Australia. There is thus an urgent need to better understand the mechanisms that can protect and promote the mental health of these individuals while adjusting to a new country. Using data from a longitudinal study of Farsi speaking refugees and asylum-seekers in Australia over a 2-year period, we hypothesized that greater post-displacement stressors measured at the onset of the study would predict lower level of physical activity and higher level of sedentary behavior at one year follow-up, which would then predict greater symptoms of depression and PTSD and lower personal mastery at two-year follow-up. Consistent with the extent literature (Hou et al., 2020), we also hypothesized that post-displacement stressors would be positively associated with symptoms of depression and PTSD and negatively associated with personal mastery at two-year follow-up.

## Methods

### Participants and study design

The study used data from the baseline, one-year and two-year follow-up assessments of the ReAssure Study. The baseline data was collected in 2017 with one-year follow-up in 2018 and two-year follow-up in 2019. The ReAssure study included participants who spoke Farsi or Dari, identified Afghanistan or Iran as their country of birth and had arrived in Australia after 2010. Participants were recruited using a representative multistage time by location sampling strategy across 16 ethno-specific grocery stores randomly selected from all known stores in Sydney, Australia. These stores are commonly visited by recently arrived Iranian and Afghan community members to purchase traditional foods, providing access to a broad cross-section of the target population and reducing recruitment bias. Customers were approached according to their time of entry, with recruitment times randomized using probability proportional to store size. For consenting households, a Kish grid (Kish, 1949) was used to randomly select one eligible adult family member for participation. Those who did not meet the inclusion criteria or those who were unable to provide informed consent were excluded. Data was collected via interviews conducted either face-to-face or by phone by bicultural researchers fluent in Farsi/Dari. Participants received AUD 20 for completing the baseline interview. Ethics approval was received by the University of New South Wales Human Research Ethics Committee (HC16637). The current study included data from those who sought refuge in Australia to capture the experiences of people from refugee and asylum-seeking backgrounds.

### Measures

#### Post-displacement stressors

An adapted version of Post-migration Living Difficulties Checklist (Steel et al., 1999) was used to assess post-displacement stressors. Participants were asked to indicate how much each of 13 presented items (e.g., not enough money to buy food, pay the rent or buy necessary clothes, discrimination, separation from family, and isolation) had been a problem for them in the past four weeks, rated on a 5-point Scale (0 =was not a problem/did not happen, 4 = a very serious problem). Items with a score of 2 (moderately serious problem) or higher were considered positive responses. A total score summing the positive responses was calculated.

#### Physical activity

The five item Simple Physical Activity Questionnaire (SIMPAQ) was used to assess the physical activity levels of participants per week (Rosenbaum et al., 2020). Participants reported the types of exercise or sports they had undertaken during the previous seven days and the duration of each activity, from which total weekly MVPA was calculated. Sedentary time was estimated using the recommended alternative SIMPAQ scoring method, which accounts for the tendency to underreport sedentary behaviour. Reported time spent in bed, walking, exercising and undertaking incidental activity was summed and subtracted from 24 hours, with the remaining time classified as sedentary.

#### Mental health outcomes

Symptoms of depression were measured using the 15-item depression subscale of the Hopkins Symptom Checklist. Each item is rated on a 4-point scale ranging from 1 (not at all) to 4 (extremely). The mean score across the 15 items was calculated to indicate depression symptom severity (McDonald’s ω = .97). PTSD symptoms were measured using the 20-item Harvard Trauma Questionnaire (HTQ) (Mollica et al., 1992) which assesses symptoms based on the DSM-IV criteria for PTSD, including recurrent thoughts and memories and feeling as though the traumatic event is happening again. Items are rated on a 4-point scale ranging from 1 (not at all) to 4 (extremely). The mean score across all items was calculated to indicate PTSD symptom severity (McDonald’s ω = .98). Personal mastery was measured using seven items developed by (Pearlin & Schooler, 1978), assessing the extent to which participants perceived themselves as having control over and being able to manage things in their lives. Items were rated on a 4-point scale (1 = strongly agree, 4 = strongly disagree) (McDonald’s ω = .97). Higher scores on these scales indicate greater levels of depression and PTSD symptoms and higher personal mastery.

#### Experiences of traumatic events

The Part-A of the Harvard Trauma Questionnaire (Mollica et al., 1992) was used to assess participants’ experiences of traumatic events, such as torture, the death of a family member, physical assault, and witnessing murder. The measure consists of 16 items rated as yes (1) or no (0). A total score, representing the number of distinct traumatic events participants indicated they had either experienced or witnessed, was calculated to indicate participants’ cumulative exposure to traumatic events.

#### Demographic variables

Key demographic information such as age, gender, education level, length of time spent in Australia was collected.

### Data analysis

We used longitudinal path analysis in R using the lavaan package to test whether engaging in moderate-to-vigorous physical activity (MVPA) and sedentary behaviours mediated the associations between post-displacement stressors and psychosocial outcomes among refugees and asylum-seekers. Post-displacement stressors at the baseline (T1) were modelled as the predictor with MVPA and sedentary behaviour at 12-month follow-up as mediators (T2), and personal mastery, depression symptoms, and posttraumatic stress disorder (PTSD) symptoms at 24-month follow-up (T3) as outcomes.

Given the moderate to strong correlation between MVPA and sedentary behaviours (r = −.55), their conceptual differences (Owen et al., 2010), and our aim to investigate their individual roles in the hypothesized relationships, two mediation models were tested. First, we examined whether MVPA mediated the associations between post-displacement stressors and each mental health outcome. Second, we examined whether sedentary behaviour mediated these associations. Direct paths were estimated from post-displacement stressors to MVPA and sedentary behavior, and from MVPA and sedentary behavior to symptoms of depression and PTSD and personal mastery. Indirect paths from post-displacement stressors to each outcome via MVPA and sedentary behavior were also estimated. We further conducted sensitivity analysis testing the same hypothesized relationships in the completers-only sample with participants who completed both baseline and 2-year follow-up assessment.

The models were controlled for age, gender, trauma and baseline levels of depression and posttraumatic stress symptoms. The significance of indirect effects was tested using bootstrapping with 5000 resamples. Full information maximum likelihood estimation was employed to use all the available information in the data. Standardized estimates, standard errors, and 95% confidence intervals are reported in Table 1.

**Table 1.** Standardized direct and indirect paths with moderate-to-vigorous physical activity.

| | $\beta$ (SE) | 95%CI<sup>a</sup><br>LLCI | ULCI |
| --- | --- | --- | --- |
| **Direct paths** |  |  |  |
| PDS → MVPA | -0.18 (0.06) | -0.292 | -0.056 |
| MVPA → Mastery | 0.19 (0.08) | 0.033 | 0.327 |
| MVPA → DEP | -0.21 (0.07) | -0.353 | -0.060 |
| MVPA → PTSDS | -0.16 (0.08) | -0.313 | 0.001 |
| PDS → Mastery | -0.34 (0.07) | -0.477 | -0.197 |
| PDS → DEP | 0.27(0.07) | 0.130 | 0.414 |
| PDS → PTSDS | 0.27 (0.07) | 0.125 | 0.408 |
| **Indirect paths** |  |  |  |
| PDS → MVPA → Mastery | -0.03 (0.02) | -0.69 | -0.003 |
| PDS → MVPA → DEP | 0.04 (0.02) | 0.006 | 0.080 |
| PDS → MVPA → PTSDS | 0.03(0.02) | -0.001 | 0.070 |

<sup>a</sup>Confidence intervals of standardized results are reported. LLCI: lower-level confidence interval. ULCI: upper-level confidence interval. PDS: Post-displacement stressors. MVPA: Moderate-to-vigorous physical activity engagement. Mastery: Personal mastery level. DEP: Symptoms of Depression. PTSDS: Posttraumatic stress disorder symptoms.

**Table 2.** Standardized direct and indirect paths with sedentary behaviour.

| | $\beta$ (SE) | 95%CI<sup>a</sup><br>LLCI | ULCI |
| --- | --- | --- | --- |
| **Direct paths** |  |  |  |
| PDS → SedBeh | 0.13 (0.05) | 0.011 | 0.243 |
| SedBeh → Mastery | -0.18 (0.08) | -0.336 | -0.011 |
| SedBeh → DEP | 0.21 (0.08) | 0.060 | 0.359 |
| SedBeh → PTSDS | 0.24 (0.08) | 0.096 | 0.386 |
| PDS → Mastery | -0.35 (0.07) | -0.485 | -0.204 |
| PDS → DEP | 0.28 (0.08) | 0.135 | 0.424 |
| PDS → PTSDS | 0.26 (0.07) | 0.117 | 0.398 |
| **Indirect paths** |  |  |  |
| PDS → SedBeh → Mastery | -0.02 (0.02) | -0.059 | 0.001 |
| PDS → SedBeh → DEP | 0.03 (0.02) | 0.001 | 0.064 |
| PDS → SedBeh → PTSDS | 0.03 (0.02) | 0.002 | 0.071 |

<sup>a</sup>Confidence intervals of standardized results are reported. LLCI: lower-level confidence interval. ULCI: upper-level confidence interval. PDS: Post-displacement stressors. SedBeh: Sedentary behavior. Mastery: Personal mastery level. DEP: Symptoms of Depression. PTSDS: Posttraumatic stress disorder symptoms.

## Results

### Participants

The sample consisted of 343 participants (80 female, 23.3%) with an average age of 35.46 (SD = 10.49) and 7.37 years in Australia (SD = 1.34). Majority (73.2%) finished high school and were single (50.7%). At the beginning of the study, more than half of the sample had insecure visa status (58%). Of 343 participants, 240 completed the MVPA and sedentary behavior at one-year follow-up (69.97%) and 192 (55.98%) completed mental health outcomes at two-year follow-up. There were no significant differences between participants who completed both T1 and T3 and those who completed only T1 in terms of gender (χ2(1) = .42, *p* = .552), age (t(332) = −1.94, *p* = .054), baseline symptoms of depression (t(332) = −.31 *p* = .754), PTSD (t(332) = −.28, *p* = .783), post-displacement stressors (t(332) = .95, *p* =.341), or number of traumatic experiences (t(332) = .02, *p* = .988).

### Longitudinal path analysis

The model fit for both moderate-to-vigorous physical activity (MVPA) and sedentary behavior was acceptable (CFI = 0.980, RMSEA= 0.068, SRMR= 0.045 for MVPA, CFI = 0.984, RMSEA = 0.067, SRMR = 0.045 for sedentary behavior).

Figures 1 and 2 depict significant standardized direct effects. Post-displacement stressors at T1 were significantly associated with lower MVPA at T1, β = −0.18, 95% CI [−0.292, −0.056] and greater sedentary behavior at T1, β = 0.13, 95% CI [0.011, 0.243].

**Figure 1.**
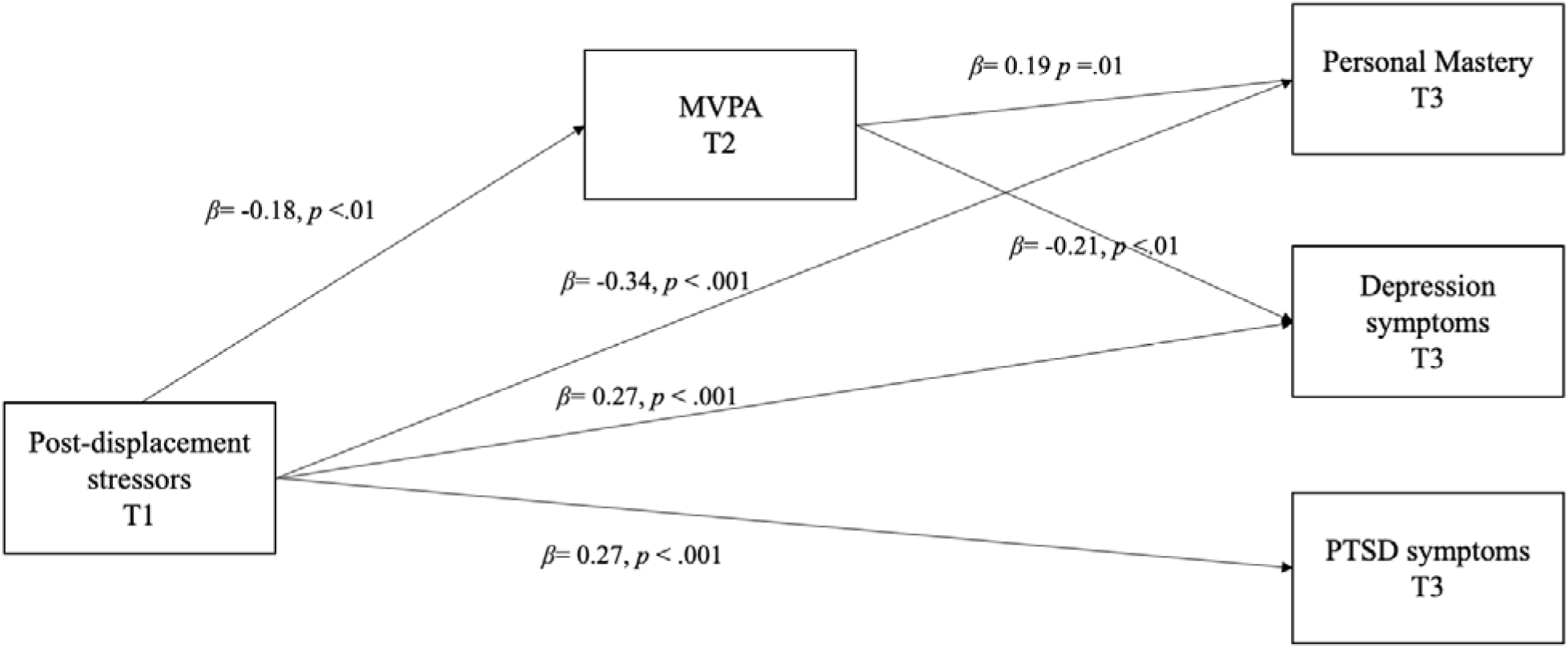
Standardized direct effects for MVPA Controlled for age, gender, traumatic experiences, and baseline levels of depression and PTSD symptoms. Only gender was significant for personal mastery (β *=* −0.14, 95% CI [−0.265, −0.024] and PTSD symptoms (β *=* 0.16, 95% CI [0.033, 0.285]. Covariances between all T3 variables were significant.

**Figure 2.**
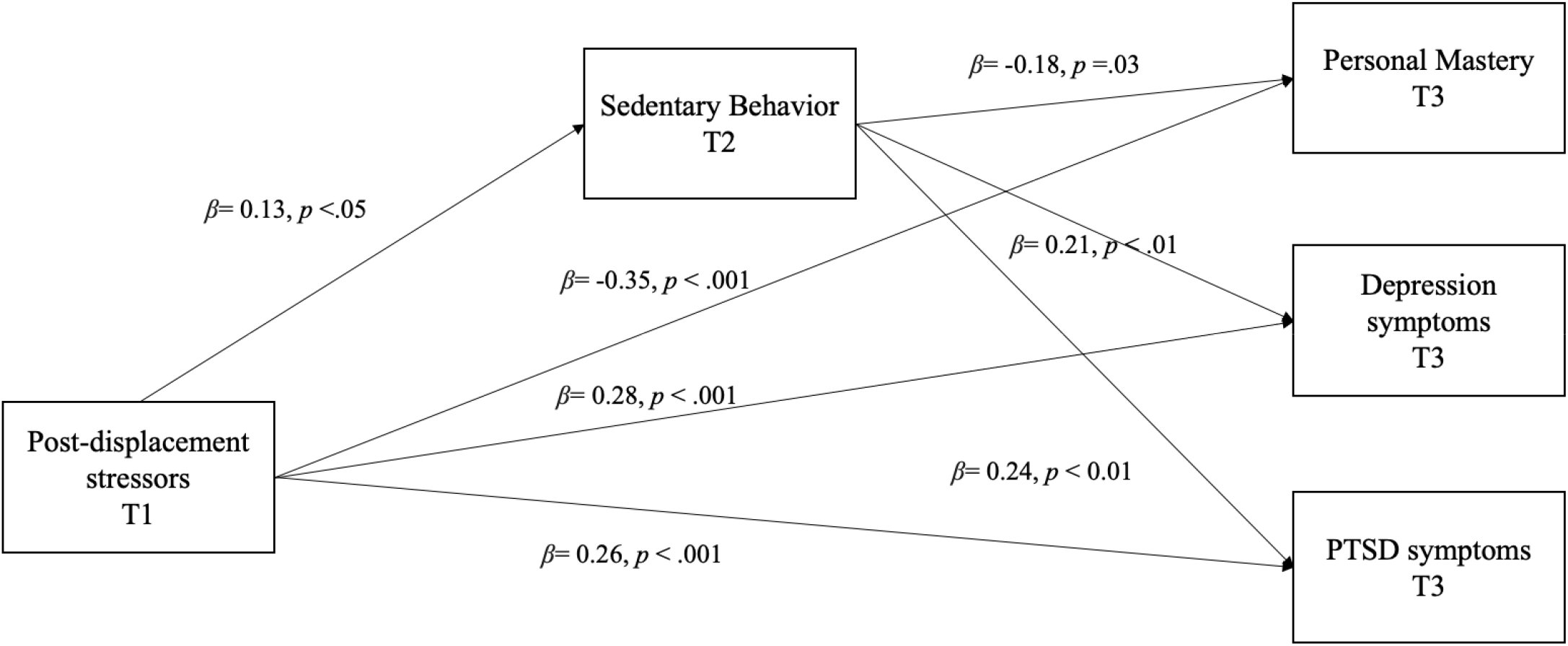
Standardized direct effects for Sedentary Behaviour. Controlled for age, gender, traumatic experiences, and baseline levels of depression and PTSD symptoms. None of the covariates were significant in the final model. Covariances between all T3 were significant.

MVPA at T2 predicted higher personal mastery, β = 0.19, 95% CI [0.033, 0.327], and lower depressive symptoms at T3, β = −0.21, 95% CI [−0.353, −0.060]. The association between MVPA at T2 and posttraumatic stress symptoms at T3 was not significant, 95% CI [−0.313, 0.001]. Sedentary behavior at T2 predicted higher depressive symptoms, β = 0.21, 95% CI [0.060, 0.359], higher posttraumatic stress symptoms, β = 0.24, 95% CI [0.096, 0.386], and personal mastery, β = −0.18, 95% CI [−0.336, −0.011]).

The indirect pathway from post-displacement stressors to personal mastery via MVPA was significant, β = −0.03, 95% CI [−0.069, −0.003], as was the indirect pathway to depressive symptoms, β = 0.04, 95% CI [0.006, 0.080]. The indirect pathway to posttraumatic stress symptoms via MVPA was not significant, 95% CI [−0.001, 0.070]. For sedentary behavior, significant indirect pathways were found for depressive symptoms, β = 0.03, 95% CI [0.001, 0.064], and posttraumatic stress symptoms, β = 0.03, 95% CI [0.002, 0.071]. The indirect pathway from post-displacement stressors to personal mastery via sedentary behavior was not statistically significant, β = −0.02, 95% CI [−0.059, 0.001]. The direct associations of post-displacement stressors with the symptoms of depression and PTSD and personal mastery were significant in both models (see Table 1 and 2). Same results were obtained with the completers-only sample (Supplementary Material 1).

## Discussion

To our knowledge, this study represents the first longitudinal investigation of the role of moderate to vigorous physical activity and sedentary behavior in the relationship between post-displacement stressors and subsequent mental health outcomes among Farsi speaking refugees and asylum-seekers. Our hypotheses were largely supported, showing that both moderate-to-vigorous physical activity and time spent sedentary partially explained the association between baseline post-displacement stressors and mental health outcomes two years later, after accounting for baseline levels of depression and PTSD and key demographic characteristics. As hypothesized, there was also a significant direct pathway from post-displacement stressors to all mental health outcomes two years later.

Our findings provide the first empirical evidence that physical activity may be one potential behavioral mechanism between stressors encountered in the resettlement environment and associated mental health problems. We found that post-displacement stressors predicted lower engagement in moderate-to-vigorous physical activity and greater time spent in sedentary behavior one year later. To date, existing studies have primarily examined the association between physical activity and mental health, rather than considering physical activity within the broader resettlement context (Filippou et al., 2024; Nilsson et al., 2021). Our findings suggest that post-displacement stressors can present significant barriers to engaging in physical activity or reducing sedentary behavior. Specifically, experiencing financial hardship and living in socially isolated neighborhoods may limit access to services, recreational facilities, and safe places to be physically active. We also found that spending less time engaging in moderate-to-vigorous physical activity and more time in sedentary behavior was associated with subsequent poorer mental health outcomes. Importantly, these behaviors are also established risk factors for chronic physical health conditions, including cardiovascular disease and type 2 diabetes, all of which contribute to premature mortality (Wahid et al., 2016). Thus, these findings highlight the importance of ensuring that people from refugee and asylum-seeker background have access to safe, culturally appropriate, and inclusive opportunities to engage in physical activity, with potential benefits extending beyond mental health to broader physical health and well-being (Rosenbaum et al., 2025).

Our mediation analyses revealed some differences across mental health outcomes. Moderate-to-vigorous physical activity partially explained the associations of post-displacement stressors with subsequent depression and personal mastery, whereas sedentary behavior partially explained their associations with depression and PTSD. One possible explanation is that these behaviors operate through distinct psychological processes. Engagement in moderate-to-vigorous physical activity such as brisk walking, jogging, and exercise involves purposeful and effortful action (Bull et al., 2020; Caspersen et al., 1985) and may provide experiences of accomplishment, competence, and control, thereby supporting personal mastery while also reducing depressive symptoms through behavioral activation (Cuijpers et al., 2026). This is consistent with recent evidence from a systematic review identifying self-efficacy and mastery as mechanisms underpinning the mental health benefits of physical activity, especially for depression (White et al., 2024). The role of personal mastery may be particularly important in the context of forced displacement, where prolonged uncertainty and insecurity can severely constrain individuals’ agency and control. Increasing opportunities for physical activity that support mastery may therefore strengthen individuals’ capacity to navigate ongoing post-displacement stressors. In contrast, time spent in sedentary behavior may reflect behavioral withdrawal and reduced engagement in daily life, processes relevant to both onset and maintenance of depression and PTSD (Dimidjian et al., 2011; Ehlers & Clark, 2000). Together, these findings suggest that moderate-to-vigorous physical activity and sedentary behavior may represent distinct behavioral pathways linking post-displacement stressors to different mental health outcomes, thereby warranting consideration as complementary targets in future intervention research. This also accords with prior research showing that sedentary behavior is not simply the absence of physical activity, as people can be physically active while also spending substantial amounts of time in sedentary behavior (Bull et al., 2020; Healy et al., 2008).

The study findings should be interpreted within some limitations. First, our measure of post-displacement stressors was based on participants’ subjective perceptions of the extent to which each stressor was problematic, rather than objective indicators of stressors encountered during resettlement. As such, the measure may partly reflect participants’ psychological distress in addition to their exposure to post-displacement stressors. Second, our sample included only refugees and asylum-seekers from Farsi speaking backgrounds, therefore, limiting the generalizability of the findings to other refugee communities in Australia. Third, we measured the time spent engaging in moderate-to-vigorous physical activity and sedentary behavior using a self-reported measure. Although our measure, SIMPAQ, has demonstrated acceptable reliability and validity against an objective measure of physical activity (e.g., accelerometer derived measures) (Rosenbaum et al., 2020), it has not been validated for Farsi language or among refugee populations. Previous studies also showed that self-reported measurement of physical activity might lead to biased estimates (Prince et al., 2008, 2020). Therefore, future studies should incorporate objective measures, such as accelerometers alongside self-report measures to improve the accuracy of physical activity assessment. Finally, although the SIMPAQ provides a pragmatic measure of total moderate-to-vigorous physical activity, it does not distinguish between domains of activity (e.g., leisure, occupational, transport or household). This may be important because accumulating evidence suggests that the association between physical activity and mental health differs by domain, with leisure-time physical activity demonstrating the strongest and most consistent associations with better mental health (Teychenne et al., 2026). Future studies should therefore examine whether specific domains of physical activity differentially mediate the relationship between post-displacement stressors and mental health.

Notwithstanding these limitations, the current findings have several implications. Firstly, physical activity and sedentary behavior may represent modifiable intervention targets for improving mental health among refugees and asylum-seekers. A meta-analysis of 27 intervention studies including both refugees and immigrants showed that physical-activity based interventions can be effective in reducing psychological distress, especially depression, PTSD, and anxiety, and improving functioning and self-efficacy (Purgato et al., 2021). More recent studies including refuges also provided evidence for the benefits of physical activity interventions such as group-based exercise and sport programs for both mental and physical health, with improvements in PTSD symptoms, psychological distress and wellbeing, and cardiovascular fitness (Knappe et al., 2024; Luttenberger et al., 2024; Nilsson et al., 2026). However, none of these interventions included a targeted component to reduce sedentary behavior. Our results underscore the importance of considering sedentary behavior in addition to increasing engagement in physical activity to maximize intervention benefits. Furthermore, there is growing recognition of integrating physical activity into psychological treatments with meta-analyses showing that physical activity delivered either as a stand-alone intervention or in combination with psychological treatments, can improve mental health outcomes among both trauma-exposed and general populations (Niemeyer et al., 2025; Thomas et al., 2020). Similarly, a recent meta-analysis found that integrated physical and psychological interventions were effective in reducing PTSD symptoms among forcibly displaced populations, providing further support for integrated approaches that address both physical and mental health needs of refugee communities (Chaudhari et al., 2025). Future interventions may therefore benefit from targeting both moderate-to-vigorous physical activity and sedentary behavior while embedding these behavioral strategies within existing mental health programs and services. In designing such interventions, recent co-design work with refugees and asylum-seekers in Australia highlights the importance of ensuring that those are culturally appropriate, trauma-informed, socially connected, and responsive to practical barriers such as affordability, accessibility, safety, and opportunities for social participation (McKeon et al., 2024). This is particularly important given that our findings suggest physical activity is a modifiable, yet socio-contextually constrained, determinant of mental health among refugees and asylum-seekers.

To conclude, this study extends the literature to investigating behavioral mechanisms underpinning the relationships between post-displacement stressors and mental health among refugees and asylum-seekers. Our findings highlight the significant role of both engaging in moderate-to-vigorous physical activity and sedentary behavior in linking post-displacement stressors to mental health outcomes. Addressing these behaviors alongside post-displacement stressors may strengthen efforts to improve mental health among refugees and asylum-seekers

## Data Availability

Data available on request due to privacy/ethical restrictions. The data that support the findings of this study are available on request from the corresponding author.

## Acknowledgement

We would like to thank each individual who participated in the study and entrusted us with their experiences. The authors used ChatGPT (OpenAI) to assist with phrasing and language editing. All content was reviewed and approved by the authors.

## Disclosure statement

None.

## Funding

This project was supported by the Australian Research Council (DP160104378).

## Author contributions

G.K: conceptualization, formal analysis, writing-original draft. R.R: data curation, project administration, writing-review and editing; G.M: writing-review and editing; S.R.: writing-review and editing and supervision; J.S.: data curation, project administration, writing-review and editing; D.B: writing-review and editing; D.S: writing-review and editing; D.H.P: writing-review and editing; Z.S: funding acquisition, writing-review and editing; R.W: supervision, writing-review and editing.

## Notes

### Competing Interest Statement

The authors have declared no competing interest.

### Author Declarations

Ethics approval was received by the University of New South Wales Human Research Ethics Committee (HC16637).

