## Supplementary Material 1 for "Longitudinal associations among post-displacement stressors, physical activity, and mental health in Farsi- and Dari-speaking refugees and asylum-seekers in Australia"

**Supplementary Table 1.** Standardized direct and indirect paths with moderate-to-vigorous physical activity.

|  |  | 95%CI^a^ |  |
| --- | --- | --- | --- |
|  | *β* (SE) | LLCI | ULCI |
| **Direct paths** |  |  |  |
| PDS 🡪 MVPA | -0.19 (0.07) | -0.329 | -0.051 |
| MVPA 🡪 Mastery | 0.18 (0.07) | 0.038 | 0.319 |
| MVPA 🡪 DEP | -0.21 (0.07) | -0.344 | -0.059 |
| MVPA 🡪 PTSDS | -0.15 (0.08) | -0.297 | 0.002 |
| PDS 🡪 Mastery | -0.34 (0.07) | -0.483 | -0.186 |
| PDS 🡪 DEP | 0.27 (0.08) | 0.122 | 0.419 |
| PDS 🡪 PTSDS | 0.27 (0.07) | 0.124 | 0.413 |
| **Indirect paths** |  |  |  |
| PDS 🡪 MVPA 🡪 Mastery | -0.04 (0.02) | -0.075 | -0.004 |
| PDS 🡪 MVPA 🡪 DEP | 0.04 (0.02) | 0.005 | 0.087 |
| PDS 🡪 MVPA 🡪 PTSDS | 0.03 (0.02) | -0.001 | 0.072 |

^a^Confidence intervals of standardized results are reported. LLCI: lower-level confidence interval. ULCI: upper-level confidence interval. PDS: Post-displacement stressors. MVPA: Moderate-to-vigorous physical activity engagement. Mastery: Personal mastery level. DEP: Symptoms of Depression. PTSDS: Posttraumatic stress disorder symptoms.

**Supplementary Table 2.** Standardized direct and indirect paths with sedentary behavior.

|  |  | 95%CI^a^ |  |
| --- | --- | --- | --- |
|  | *β* (SE) | LLCI | ULCI |
| **Direct paths** |  |  |  |
| PDS 🡪 SedBeh | 0.17 (0.07) | 0.046 | 0.301 |
| SedBeh 🡪 Mastery | -0.18 (0.08) | -0.334 | -0.010 |
| SedBeh 🡪 DEP | 0.21 (0.08) | 0.059 | 0.351 |
| SedBeh 🡪 PTSDS | 0.24 (0.07) | 0.092 | 0.378 |
| PDS🡪 Mastery | -0.35 (0.08) | -0.493 | -0.195 |
| PDS 🡪 DEP | 0.28 (0.08) | 0.127 | 0.431 |
| PDS 🡪 PTSDS | 0.26 (0.07) | 0.116 | 0.406 |
| **Indirect paths** |  |  |  |
| PDS 🡪 SedBeh 🡪 Mastery | -0.03 (0.02) | -0.075 | 0.001 |
| PDS 🡪 SedBeh 🡪 DEP | 0.04 (0.02) | 0.004 | 0.079 |
| PDS 🡪 SedBeh 🡪 PTSDS | 0.04 (0.02) | 0.007 | 0.087 |

^a^Confidence intervals of standardized results are reported. LLCI: lower-level confidence interval. ULCI: upper-level confidence interval. PDS: Post-displacement stressors. SedBeh: Sedentary behavior. Mastery: Personal mastery level. DEP: Symptoms of Depression. PTSDS: Posttraumatic stress disorder symptoms.
